## Supplemental Materials for "Impact of a multi-pronged cholera intervention in an endemic setting"

### Supplementary information

#### Testing strategy

Testing rate was high: 1580 out of the 1634 (96.7%) suspected cholera cases residing in Kalemie consulting in the treatment center were tested to confirm the diagnosis by stool culture and/or PCR (Figure S1). Overall, a stool culture was performed on 1325 suspected cholera cases, and both a stool culture and a PCR were performed on 255 suspected cholera cases. Out of the 255 cases who were tested by both PCR and stool culture:107 were culture -/PCR -, 22 were culture +/PCR +, 105 were culture -/PCR +, and 21 were culture +/PCR -.

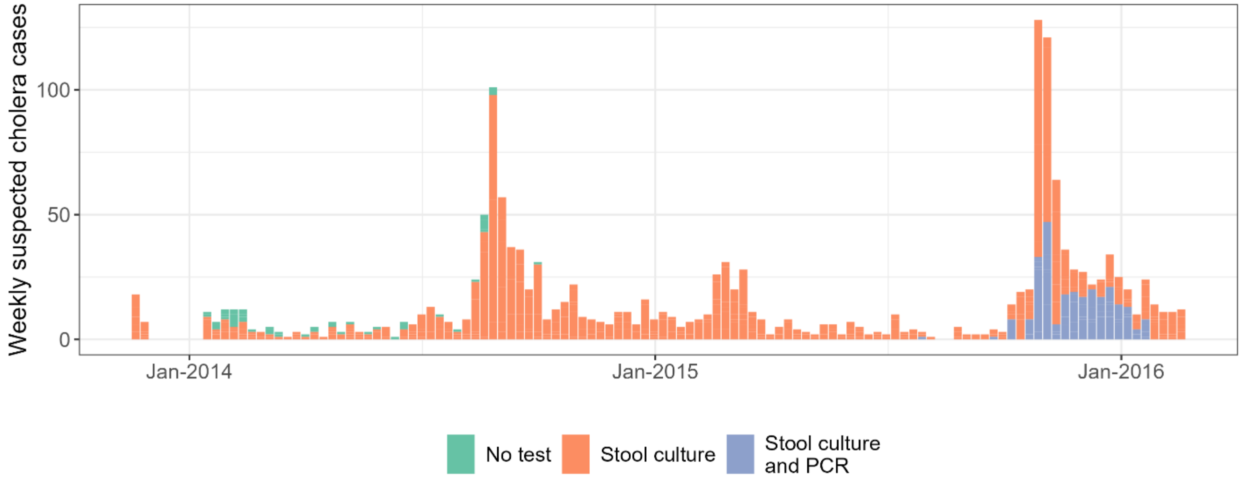


*Figure S1:Testing of the suspected cholera cases residing in Kalemie*

Stool cultures were performed at the reference national laboratory in Kinshasa after collection and enrichment on alkaline peptone water. PCR tests were performed by Pasteur Institute in Paris after collection on dry paper and enrichment on alkaline peptone water.

We did not use confirmed cholera cases to perform the analysis because of the heterogeneous testing strategy and suspicions of contamination of part of the stool cultures. Both stool cultures and PCR have very high specificity when performed in optimal conditions, but their precise diagnostic qualities to confirm cholera are hard to assess because of the lack of a “perfect” reference (1). However, PCR should be more sensitive than stool cultures because it does not require *Vibrio cholerae* (*V. cholerae*) to be alive or in a culturable state to be detected. The two phases of the testing strategy have different positive predictive value (PPV), for which we have no clear estimates. Using those different phases of laboratory confirmation to fit the model would have required making additional assumptions or adding parameters. Regarding our suspicions of contaminations, 16.4% (21/128) of the suspected cases that were PCR - were culture +. The group of 21 PCR -/culture + suggests that contaminations could have occurred, which limits the confidence on the stool culture results prior to the introduction of PCR.

Instead of relying on the number of confirmed cholera cases, the models presented in the manuscript make the assumption, through the parameter $r$, that the number of suspected cholera cases reflects a portion of the “true number of cholera cases” in the population. We further explore below if it is a reasonable assumption.

### Analysis of the stability of reporting rate over time

The meaning of $r$, the reporting proportion, can be better understood by estimating the number of true cases ($C_t$) captured by reported suspected cases ($A_t$) (PP in Table S1). Only a portion ($\rho$) of all the cholera cases ($C_t$) will go to the treatment center, because of the severity of the symptoms and/or their access to care. So, the number of the true cases that could be captured by the reported suspected cholera cases is $C_t \times \rho$ (P in Table S1). Some of them will accurately be counted as suspected cholera cases, the true positives (TP in Table S1), and some of them will not fit the suspected case definition and will not be counted as suspected cholera cases, the false negatives (FN in Table S1). The suspected cholera cases also include diarrheal diseases that are not cholera, the false positives (FP in Table S1). Some of those non-cholera diarrheal diseases will not be counted as suspected cholera cases, the true negatives (TN in Table S1).

*Table S1:* Relationships between the true cholera cases consulting and the reported suspected cases through the imperfect case definition

| | Suspected cases or predicted positives (PP=TP+FP) $A_t$ | Non suspected cases or predicted negatives (PN=FN+TN) |
|---|---|---|
| True cases consulting or just positives (P=TP+FN): $C_t \times \rho$ | True positives (TP) | False negatives (FN) |
| Non cases or just negatives (N=FP+TN) | False positives (FP) | True negatives (TN) |

The proportion of true cases consulting captured by the suspected case definition reflects the sensitivity of the case definition ($Se = TP/P$), so: $TP = C_t \times \rho \times Se$. It can also be estimated by multiplying the reported suspected cases ($A_t$) and the positive predictive value of the suspected case definition ($PPV = TP/PP$), so $TP = A_t \times PPV$.

$$C_t \times \rho \times Se = A_t \times PPV$$

It implies that $A_t = \frac{C_t \times \rho \times Se}{PPV}$ and then $r = \frac{\rho \times Se}{PPV}$

$PPV$ varies with the prevalence of the infection among people consulting at the treatment center ($prev$):

$$PPV = \frac{Se \times prev}{Se \times prev + (1 - Sp)(1 - prev)}$$

With $prev = \frac{P}{P+N}$ with $N = \frac{FP}{1-Sp} = \frac{PP-TP}{1-Sp} = \frac{(A_t - C_t \times \rho \times Se)}{(1-Sp)}$ with Sp ($Sp = TN/N$) the specificity of the case definition.

Then $prev = \frac{C_t \times \rho}{C_t \times \rho + \frac{(A_t - C_t \times \rho \times Se)}{(1-Sp)}}$ with $A_t \geq C_t \times \rho \times Se$

We explored plausible values for $prev$ using reasonable values for $\rho$, our average model predictions of $C_t$, the reported cases $A_t$, and estimates of the sensitivity (Se) and specificity (Sp) of the case definition from the literature. The DRC uses the WHO case definition for suspected cholera cases, with an estimated sensitivity of 92.7% and a specificity of 8.1% (2). Figure S2A shows the distribution of $prev$ when exploring a range of values for $\rho$ below 10%. An upper limit of 10% seems reasonable given that as many as 80% of cases are asymptomatic in endemic settings (3) and where only symptomatic and severe cases are likely to seek care. We then explored possible values for $prev$ using 20 values of $\rho$ equally spaced between 0.01 and 0.1 using the set of $C_t$ and $A_t$for each of them. The resulting estimates of $prev$ (after excluding combinations leading to $A_t < C_t \times \rho \times Se$) provide a range of values that we considered as reasonable. We used them to define the corresponding PPV and then explore $r$. The distribution of what we expect as reasonable values for $prev$ has 2.5$^{th}$ and 97.5$^{th}$ percentiles of 0.006 and 0.45 with a mean of 0.11 and a median of 0.07 (Figure S2).

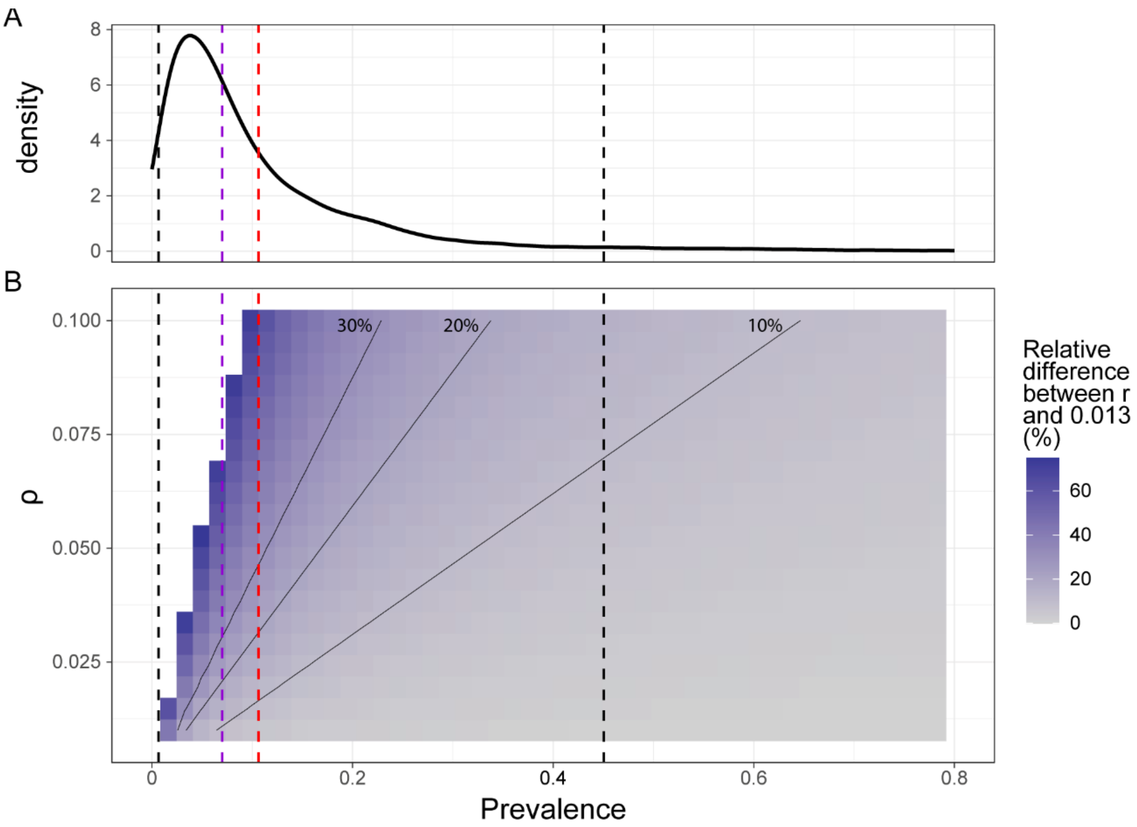


*Figure S2:* Distribution of reasonable values for prevalence and relative variation between r and our model estimate, 0.013, over a range of values for $\rho$ and prevalence. (A) Density plot of prevalence when considering a range of values for $\rho$ below 0.1, a sensitivity of 0.921 and a specificity of 0.081 (2), model predictions for $C_t$, and reported cases $A_t$. The vertical black, purple, and redlines dashed lines indicate the 2.5$^{th}$ and 97.5$^{th}$ percentiles, the median, and the mean of our estimates for prevalence. (B) Heat map of the relative difference between r and our model estimate of r (0.013) given a range of values for $\rho$ and prevalence. The thin black lines are isolines indicting relative difference of 10%, 20%, and 30%. The values of $r$ above 1 were clipped.

We then explored the sensitivity of $r$ to a range of values of $prev$ and $\rho$, with $prev$ determining the values of $PPV$. Figure S2B shows the variation of the relative difference between the resulting estimates of $r$ and our average model estimate, 0.013, with $prev$ and $\rho$. When excluding the values of $r$ above 1 and restricting the estimates corresponding to a range of $prev$ between its 2.5$^{th}$ and 97.5$^{th}$ percentiles (between the two black vertical dashed lines on Figure S2), 33.0% had a maximum of 10% relative difference and 67.0% had a maximum of 20% relative difference with 0.013.

Although it is a post hoc analysis, there appears to be a wide region of the parameter space where $r$ is relatively insensitive to fluctuations of $prev$, its main component likely to change substantially over time. This suggests that $r$ could be approximated as constant over time in some region of the parameter space.

#### Confirmed cholera cases and model prediction

We further checked how reasonable our assumption that $r$ is constant over time is by comparing the confirmed cases and our model predictions. The assumption that the reported suspected cholera cases ($A_t$) reflect the "true number of cholera cases" implies that the proportion of cholera cases it captured is roughly constant over time. If it is a reasonable assumption given the available data, our model predictions of the reported suspected cases ($A_t$) (not the observed reported cases) should be consistent with the observed confirmed cases when randomly drawing an expected number of confirmed cases ($Pos_t$) based on our model using a constant probability to be tested and found positive ($p(test \cap pos)$), with $p(test \cap pos) = p_{test} \times p_{pos}$. However, we still accounted for the change in testing strategy by having two values for $p_{pos}$.

$$Pos_t \sim Binom(A_t, p(test \cap pos))$$

$p_{test} = \frac{1580}{1634} = 0.967$, and $p_{pos} = \begin{cases} 0.107, when\ only\ stool\ cultures\ were\ done \\ 0.248, when\ both\ stool\ cultures\ and\ PCR\ were\ done \end{cases}$

Assuming that there are no false positive results, 10.7 % of the tested suspected cases were confirmed when only stool cultures were used, and 24.8% after the addition of PCR.

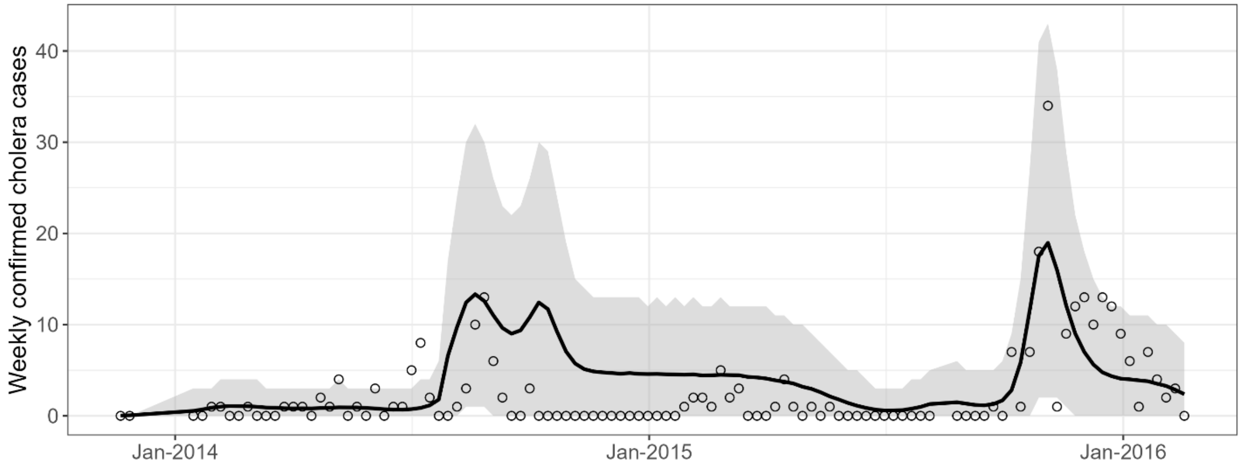


*Figure S3:* Confirmed cholera cases and expected confirmed cholera cases based on model predictions. Confirmed cholera cases are indicated with empty circles. The average expected number of cholera cases (black line and its 95% credible interval (grey envelope) were obtained by drawing from a binomial distribution with the probability to be tested and found positive $p(test \cap pos)$ over 10,000 simulated time series of reported suspected cases based on the best performing model.

Based on our model, the expected number of confirmed cholera cases using this strategy captured the observed confirmed cases well, 97.2% (106/109) of the observed weekly confirmed cases fall within the 95% credible interval (Figure S3). This good fit is not evidence that suspected cholera cases is a robust proxy for "true cholera cases", but it supports the idea that it is a reasonable proxy during this limited period of time given the data and their limitations. This also likely reflects a strong uncertainty when using confirmed cases because of their low numbers.

### Additional details on some parameters and inputs

#### Step function η

The step function η includes three components:

- An estimate of the vaccine coverage at the level of the city of Kalemie
- An estimate of vaccine effectiveness
- A penalty to account for the spatially targeted nature of the vaccination campaign

Drawing conclusions on the individual values of the vaccine effectiveness and the penalty term is not possible based on our model fits because of the product of those parameters. However, considering reasonable values for each of them allows us to screen a range of meaningful values for η in our models (see model selection below).

##### Vaccine coverage

Doctors Without Borders (*Médecins Sans Frontières*, MSF) performed a vaccination coverage survey in August 2014 (4). They shared the estimated vaccine coverage at the health area level for participants who received 1 or 2 doses as in July 2014. We estimated the vaccine coverage at the city level by adding the products of the estimates of population sizes and vaccine coverage at the same geographical level and dividing it by the total population size.

$$VC_1 = \frac{\sum VC_1^n \times N^n}{\sum N^n} \text{ and } VC_2 = \frac{\sum VC_2^n \times N^n}{\sum N^n}$$

The estimated vaccine coverage, $VC_1$, for November 2013 is simply the number of distributed doses divided by the population of Kalemie. Since only 1 dose was distributed for 3 days this avoids the inherent imprecision of estimating vaccine coverage retrospectively, that cannot be avoided when multiple doses were distributed, like in July 2014.

MSF also performed a census in the vaccine targeted areas of Kalemie city. We used those estimates and estimated the population size of the remaining areas as the difference between those areas and Kalemie's estimated population size (262,963).

Few people received three doses. This would have occurred either because they received one dose during the 3 days doses were distributed in November 2013 and two doses in July 2014 or they received two doses in July 2014 and also benefitted from the short catch-up done in August 2014 to provide a dose to people who could not receive the first dose. We did not consider that receiving three doses provided a higher protection than two doses, because it was anecdotal.

Susceptible individuals receiving a vaccine moved to the recovered compartment (with a probability corresponding the vaccine effectiveness) 1 week after receiving the dose(s). This was done 1 week after receiving the first dose for people receiving two doses but with a probability corresponding to the VE of a two-dose regimen.

##### Vaccine effectiveness

Estimates of short-term vaccine effectiveness (VE) of Shanchol$^{TM}$ are scarce. There are estimates made in the aftermath of reactive vaccination campaigns in Zambia and Guinea using laboratory confirmed symptomatic cases and controls sampled from the population without recent cholera-like symptoms, respectively 88.9% for one dose and 86.6% for two doses (5,6). However, these estimates are not precise (relatively large 95% confidence intervals) and likely overestimate the effectiveness to prevent an infection (symptomatic or not).

Overall, we considered three sets of assumptions regarding VE in addition to the estimates from Zambia and Guinea, during our model selection (Table S2).

*Table S2 :* Vaccine effectiveness assumptions used in the model selection

| Vaccine effectiveness assumption | Vaccine effectiveness for 1 dose (%) | Vaccine effectiveness for 2 doses (%) |
|---|---|---|
| 1 | 88.9 | 86.6 |
| 2 | 80 | 85 |
| 3 | 70 | 75 |
| 4 | 60 | 65 |

##### Penalty term

The penalty term is a way to account for the higher level of immunity in the targeted areas due to higher exposure to cholera historically. We considered a set of different values during our model selection: 0.7, 0.8, 0.9, and 1 (no higher immunity in the targeted areas).

#### Seasonal migration

Nighttime radiance has been shown to be a good proxy for human presence and it has been used to estimate population size variation in high and low-income countries (7–11). Its variation has previously been used to infer seasonal population variation and to account for it in Susceptible Infected Recovered models (12).

We extracted nighttime radiance data from Visible Infrared Imaging Radiometer (13) data from January 1st 2012 until December 31st 2018 in the Kalemie city using georeferenced polygons of the boundaries of the city of Kalemie created by MSF.

Radiance data from nights with heavy cloud cover or within 10 days of a full moon were excluded to avoid artificially low or high radiance values. We calculated a 7-day moving average, counting the number of data points used to calculate it, and extracted the seasonal pattern by fitting a Generalized Additive Model (GAM) with a weighted cyclical spline, the weights being the number of data points used to calculated the averages. The seasonal variation of the population size was then estimated using the 1st derivative of the cyclical spline ($rad_t$) assuming a linear relationship.

$$f_S(rad_t) = \alpha_1 rad_t$$

$$f_I(rad_t) = \alpha_I rad_t$$

With

$$ratio_{S/I} = \frac{\alpha_1}{\alpha_I}$$

We explored a set of assumptions regarding the ratio of susceptible and infected individuals among the migrating population ($ratio_{S/I}$): 10, 40, 70, and 100. The assumptions about $ratio_{S/I}$ systematically favor susceptible individuals since symptomatic infected individuals are less likely to travel. We did not estimate the number of recovered individuals in the migrating population because they do not directly contribute to transmission.

#### Environmental drivers

We considered three main environmental drivers influencing *V. cholerae* abundance in the environment, exposure to the environmental reservoir, or contamination of the environment:

- Lake surface temperature of the lake water surrounding the city of Kalemie
- Chlorophyll-a content in the lake water surrounding the city of Kalemie
- Rainfall in the area around the city of Kalemie

##### Lake surface temperature

We extracted daily lake surface temperatures (in Celsius degrees) from Moderate Resolution Imaging Spectrometer (MODIS) (14). We only used the values from the raster cells whose centroids fell within a 3 km buffer around Kalemie city during the whole study period (from November 2013 to February 2016).

We interpolated the missing values by calculating a 7-day moving average and counting the number of data points used to calculate it. We then smoothed the 7-day averages with a weighted loess regression, using the number of data points as weights. The values used as input in the models are the smoothed values after standardization.

##### Chlorophyll-a

We extracted daily near surface chlorophyll-a concentration (in $mg/m^3$) in the lake water from MODIS (14). The estimates available over time are patchy for chlorophyll-a, so we minimized the number of missing values by expanding the buffer around the city of Kalemie. We extracted the values from the raster cells whose centroids fell within a 30 km buffer during the whole study period (from November 2013 to February 2016). We do not expect over/under-estimating the variation of chlorophyll-a in the surroundings of Kalemie because it has been shown to fluctuate reasonably similarly within wide coherent ecological regions bigger than our buffer (15).

As for lake surface temperature, we interpolated the missing values by calculating a 7-day moving average and counting the number of data points used to calculate it. We then smoothed the 7-day averages with a weighted loess regression, using the number of data points as weights. The values used as input in the models are the smoothed values after standardization.

##### Rainfall

We extracted daily precipitation estimates (in $kg/m^2/s$) around Kalemie from meteorological forcings data (16). We only used the values from the raster cells whose centroids fell within a 20 km buffer around Kalemie city during the whole study period (from November 2013 to February 2016) .

We calculated a 7-day moving average and smoothed it with a loess regression. The values used as input in the models for $f(rain_t)$ are the smoothed values after normalization.

The magnifying effect of rainfall on environmental exposure or environmental contamination has been explored in an epidemic setting in South Sudan (17) in the form of $\left(1+\lambda\left(\frac{rain_t}{\max(rain_t)}\right)^{\alpha}\right)$. To minimize identifiability issues between $\lambda$ and $\alpha$, we simplified the expression and assumed a linear relationship ($\alpha$=1).

### Model selection and model fit

Every model ran on 3.7 GHz 4th generation AMD EPYC processors. Getting 300,000 samples took about 12 days.

We had a burn-in period of 200 thousand samples and then saved 100 thousand samples from the posterior distribution.

We assessed the fit of every candidate model with visual inspection of the average prediction and by calculating the widely applicable information criterion (WAIC) (18).

We first fit a group of 96 models (Table S3):

- 64 "full" models with migration of susceptible and infected individuals using every combination of the assumptions for VE, the penalty Θ, and $ratio_{S/I}$.
- 16 models with only susceptible individuals migrating ($f_I(rad_t)=0$) using every combination of assumptions for VE and the penalty Θ
- 16 models without migrating population ($f_S(rad_t)=0$ and $f_I(rad_t)=0$) using every combination of assumptions for VE and the penalty Θ. The model presented in the manuscript had the lowest WAIC among the models with fewer parameters. It has a penalty Θ of 0.8, a VE of 60% and 65% for 1 and 2 doses (vaccine effectiveness assumption 4) (in bold in Table S3).

We selected the "best performing model" as described in the manuscript and fit it with alternative structure as sensitivity analysis:

- 1 model without environmental compartment relying only on direct interhuman transmission ($\beta_e=0$)
- 1 model without bacterial growth: the net bacterial growth is constant and negative without fluctuation based on environmental input ($\varphi_t=0$)

The 2 models without environment compartment or bacterial growth are simpler models used to demonstrate that those two elements are necessary to reproduce the cholera transmission observed over the 118 week-study period.

#### Varying assumptions for vaccine effectiveness, penalty term, and $\text{ratio}_{S/I}$

None of the 96 models were significantly different in terms of WAIC (Figure S4, and Table S3). The model presented in the manuscript was chosen because it had the lowest WAIC among the models with the fewest number of parameters (model without seasonal migration).

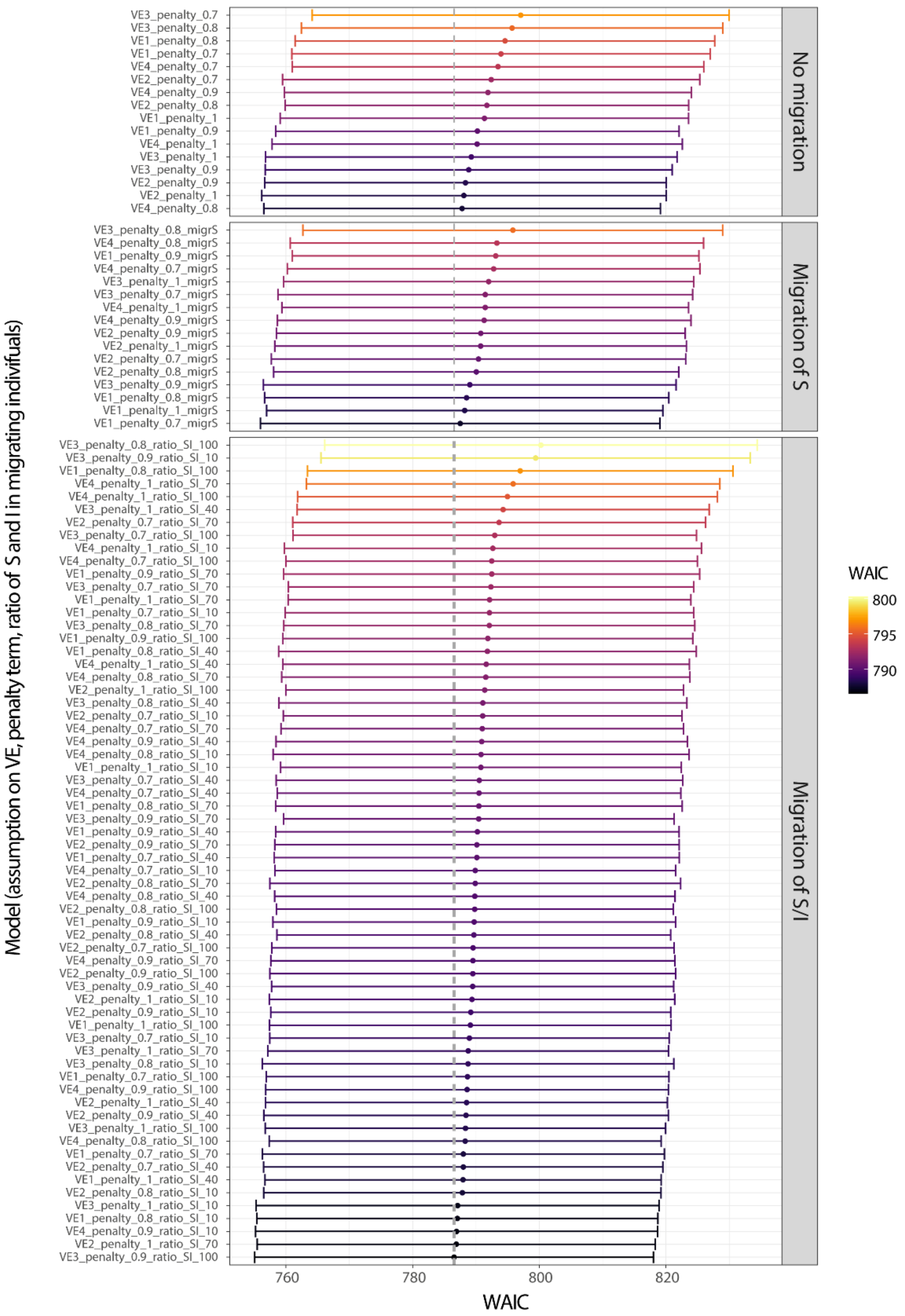


*Figure S4:* Model fit of the 96 models assessed by WAIC. The error bars provide a range of 1.96 standard errors around the WAIC and provide a visual guide on whether or not the difference in WAIC is significant. The vertical dashed grey line indicates the minimal value of WAIC across the 96 models. The names of the models indicate 1) the assumption on the vaccine effectiveness values, the penalty term ($\Theta$) and the $ratio_{S/I}$ for "full" models, or migrS for models with only susceptible individuals migrating, or nothing for no migration.

*Table S3:*Model fit of the 96 models explored with every combination of the assumptions on vaccine effectiveness, penalty term, and ratio of susceptible over infected individuals for the models with migrating population[a]

| Type of model | Vaccine effectiveness assumptions | Penalty term | $ratio_{S/I}$ | WAIC | Difference in WAIC[b] | WAIC standard error |
|---|---|---|---|---|---|---|
| "Full" model with migration of susceptible and infected individuals | 1 | 0.7 | 10 | 792.1 | 5.6 | 32.2 |
| | 1 | 0.7 | 40 | 790.1 | 3.6 | 31.9 |
| | 1 | 0.7 | 70 | 788 | 1.5 | 31.7 |
| | 1 | 0.7 | 100 | 788.7 | 2.1 | 31.8 |
| | 1 | 0.8 | 10 | 787.1 | 0.5 | 31.6 |
| | 1 | 0.8 | 40 | 791.8 | 5.3 | 33 |
| | 1 | 0.8 | 70 | 790.5 | 3.9 | 32.1 |
| | 1 | 0.8 | 100 | 797 | 10.4 | 33.6 |
| | 1 | 0.9 | 10 | 789.7 | 3.2 | 31.8 |
| | 1 | 0.9 | 40 | 790.2 | 3.7 | 31.8 |
| | 1 | 0.9 | 70 | 792.5 | 5.9 | 32.8 |
| | 1 | 0.9 | 100 | 791.9 | 5.3 | 32.3 |
| | 1 | 1 | 10 | 790.8 | 4.2 | 31.6 |
| | 1 | 1 | 40 | 788 | 1.4 | 31.3 |
| | 1 | 1 | 70 | 792.1 | 5.6 | 31.8 |
| | 1 | 1 | 100 | 789.1 | 2.6 | 31.7 |
| | 2 | 0.7 | 10 | 791.1 | 4.5 | 31.5 |
| | 2 | 0.7 | 40 | 788 | 1.4 | 31.5 |
| | 2 | 0.7 | 70 | 793.7 | 7.1 | 32.6 |
| | 2 | 0.7 | 100 | 789.5 | 3 | 31.7 |
| | 2 | 0.8 | 10 | 787.9 | 1.3 | 31.4 |
| | 2 | 0.8 | 40 | 789.7 | 3.1 | 31.1 |
| | 2 | 0.8 | 70 | 789.9 | 3.3 | 32.4 |
| | 2 | 0.8 | 100 | 789.8 | 3.3 | 31.3 |
| | 2 | 0.9 | 10 | 789.2 | 2.6 | 31.6 |
| | 2 | 0.9 | 40 | 788.4 | 1.9 | 31.9 |
| | 2 | 0.9 | 70 | 790.2 | 3.6 | 31.9 |
| | 2 | 0.9 | 100 | 789.5 | 3 | 32 |
| | 2 | 1 | 10 | 789.4 | 2.8 | 32 |
| | 2 | 1 | 40 | 788.5 | 2 | 31.7 |
| | 2 | 1 | 70 | 786.9 | 0.3 | 31.4 |
| | 2 | 1 | 100 | 791.4 | 4.8 | 31.4 |
| | 3 | 0.7 | 10 | 789 | 2.4 | 31.5 |
| | 3 | 0.7 | 40 | 790.5 | 4 | 32.1 |
| | 3 | 0.7 | 70 | 792.4 | 5.8 | 32 |
| | 3 | 0.7 | 100 | 793 | 6.4 | 31.9 |
| | 3 | 0.8 | 10 | 788.8 | 2.2 | 32.5 |
| | 3 | 0.8 | 40 | 791.1 | 4.5 | 32.2 |
| | 3 | 0.8 | 70 | 792.1 | 5.6 | 32.4 |
| | 3 | 0.8 | 100 | 800.3 | 13.7 | 34.1 |
| | 3 | 0.9 | 10 | 799.4 | 12.9 | 33.9 |
| | 3 | 0.9 | 40 | 789.5 | 2.9 | 31.7 |
| | 3 | 0.9 | 70 | 790.5 | 3.9 | 30.8 |
| | **3** | 0.9 | 100 | 786.5 | 0 | 31.5 |
| | 3 | 1 | 10 | 787.1 | 0.6 | 31.8 |
| | 3 | 1 | 40 | 794.3 | 7.8 | 32.5 |
| | 3 | 1 | 70 | 788.8 | 2.2 | 31.6 |

| | | | | | | |
|---|---|---|---|---|---|---|
| | 3 | 1 | 100 | 788.3 | 1.8 | 31.6 |
| | 4 | 0.7 | 10 | 789.9 | 3.4 | 31.6 |
| | 4 | 0.7 | 40 | 790.5 | 3.9 | 31.8 |
| | 4 | 0.7 | 70 | 791 | 4.4 | 31.8 |
| | 4 | 0.7 | 100 | 792.5 | 5.9 | 32.5 |
| | 4 | 0.8 | 10 | 790.8 | 4.3 | 32.8 |
| | 4 | 0.8 | 40 | 789.8 | 3.3 | 31.6 |
| | 4 | 0.8 | 70 | 791.6 | 5 | 32.2 |
| | 4 | 0.8 | 100 | 788.3 | 1.7 | 30.9 |
| | 4 | 0.9 | 10 | 786.9 | 0.4 | 31.7 |
| | 4 | 0.9 | 40 | 790.9 | 4.4 | 32.5 |
| | 4 | 0.9 | 70 | 789.5 | 3 | 31.9 |
| | 4 | 0.9 | 100 | 788.6 | 2.1 | 31.8 |
| | 4 | 1 | 10 | 792.7 | 6.1 | 32.9 |
| | 4 | 1 | 40 | 791.6 | 5.1 | 32.1 |
| | 4 | 1 | 70 | 795.9 | 9.3 | 32.6 |
| | 4 | 1 | 100 | 795 | 8.4 | 33.1 |
| **Only susceptible individuals migrating** | 1 | 0.7 | | 787.5 | 1 | 31.5 |
| | 1 | 0.8 | | 788.5 | 2 | 31.9 |
| | 1 | 0.9 | | 793.1 | 6.6 | 32.1 |
| | 1 | 1 | | 788.2 | 1.7 | 31.3 |
| | 2 | 0.7 | | 790.4 | 3.9 | 32.7 |
| | 2 | 0.8 | | 790 | 3.5 | 32 |
| | 2 | 0.9 | | 790.8 | 4.2 | 32.3 |
| | 2 | 1 | | 790.7 | 4.2 | 32.5 |
| | 3 | 0.7 | | 791.5 | 4.9 | 32.7 |
| | 3 | 0.8 | | 795.8 | 9.3 | 33.1 |
| | 3 | 0.9 | | 789 | 2.5 | 32.6 |
| | 3 | 1 | | 792 | 5.5 | 32.4 |
| | 4 | 0.7 | | 792.8 | 6.2 | 32.5 |
| | 4 | 0.8 | | 793.3 | 6.8 | 32.6 |
| | 4 | 0.9 | | 791.3 | 4.8 | 32.6 |
| | 4 | 1 | | 791.5 | 4.9 | 32.1 |
| **No migration** | 1 | 0.7 | | 794 | 7.4 | 33 |
| | 1 | 0.8 | | 794.6 | 8 | 33.1 |
| | 1 | 0.9 | | 790.2 | 3.7 | 31.8 |
| | 1 | 1 | | 791.3 | 4.8 | 32.2 |
| | 2 | 0.7 | | 792.4 | 5.9 | 32.9 |
| | 2 | 0.8 | | 791.7 | 5.2 | 31.8 |
| | 2 | 0.9 | | 788.3 | 1.8 | 31.7 |
| | 2 | 1 | | 788.1 | 1.6 | 31.9 |
| | 3 | 0.7 | | 797.1 | 10.5 | 32.9 |
| | 3 | 0.8 | | 795.7 | 9.2 | 33.3 |
| | 3 | 0.9 | | 788.9 | 2.3 | 32.1 |
| | 3 | 1 | | 789.3 | 2.7 | 32.5 |
| | 4 | 0.7 | | 793.5 | 6.9 | 32.5 |
| | **4** | **0.8** | | **787.8** | **1.3** | **31.3** |
| | 4 | 0.9 | | 791.9 | 5.3 | 32.1 |
| | 4 | 1 | | 790.2 | 3.7 | 32.4 |

a: The model presented in the manuscript is in bold.

b: The difference in WAIC is calculated with the model with the lowest value as a reference.

#### Sensitivity analysis: different model structure

Removing the environmental reservoir or removing the bacterial growth based on environmental inputs, implying that net bacterial growth cannot become positive, significantly decreased model fit (WAIC of 787.8, 853.6, and 868.8 respectively for VE4_penalty_0.8, no environmental reservoir, and no bacterial growth) (Figure S5).

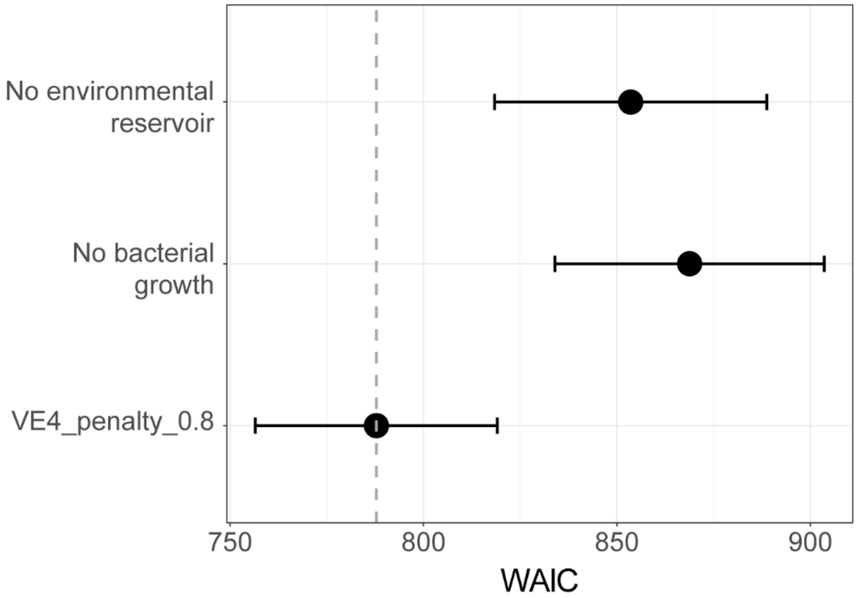


*Figure S5:* Comparison of the model fit of the model the lowest WAIC among the models with the lowest number of parameters and its simpler variations: one without environmental reservoir and one without bacterial growth. The full circles indicate the corresponding WAIC and the error bars provide a range of 1.96 standard errors around the WAIC. The vertical dashed grey line indicates the minimal value of WAIC.

### Assessing convergence

We assessed convergence visually and through the Gelman and Rubin statistic (Rhat). Rhat is commonly interpreted as indicating convergence when below 1.1. We ran two chains for the main model with two different starting values. The starting values for the first chain were chosen following an iterative process when fitting the model. The second set of starting values was chosen to intentionally start sampling the posterior distribution in an area of the parameter space that favored an interpretation of the main drivers of transmission that would differ from the one provided by the first chain: a transmission more dominated by its direct interhuman component in a population that is primarily susceptible. We explored alternative starting values reflecting scenarios even further away from the interpretation provided by the first chain and also modified the parameters linked to bacterial growth. Those parameters appeared sensitive enough to lead to chains that would not converge after 300,000 samples when using values too different from the ones used in the two chains presented here. Due to the computational cost, we decided not to add more chains.

We assessed convergence in three steps:

- we looked at the Rhat, the posterior distribution, and Monte-Carlo standard error of the individual parameters
- we looked at the Rhat and the posterior distribution of groups of parameters to further explore potential identifiability issues
- we looked at the Rhat and the posterior distribution of primary indicators of interest to assess the impact of the intervention: number of infected individuals and net bacterial growth

#### Individual parameters

*Table S4:* Prior distribution and starting values used for the two chains and corresponding Gelman and Rubin statistics (Rhat)

| Parameter | Priors | Chain 1-Starting values | Chain 2-Starting values | Rhat |
|---|---|---|---|---|
| $\boldsymbol{log(\beta_h)}$ | Normal distribution with<br>• Mean=0<br>• Standard deviation=0.5 | -0.34 | 0.25 | 1.000 |
| $\boldsymbol{\beta_e}$ | Truncated normal distribution with<br>• Mean=0<br>• Standard deviation=0.5<br>• Left-truncated at 0 | 0.36 | 0.025 | 1.031 |
| $\boldsymbol{\beta_{WASH}}$ | Truncated normal distribution with<br>• Mean=0<br>• Standard deviation=0.5<br>• Left-truncated at 0<br>• Right-truncated at 2 | 0.06 | 0 | 1.006 |
| $\boldsymbol{\alpha_2}$ | Normal distribution with<br>• Mean=0<br>• Standard deviation=0.5 | 0.33 | 0.28 | 1.041 |
| $\boldsymbol{\alpha_3}$ | Normal distribution with<br>• Mean=0<br>• Standard deviation=0.5 | 0.009 | 0.009 | 1.000 |
| $\boldsymbol{\alpha_4}$ | Normal distribution with<br>• Mean=0<br>• Standard deviation=0.5 | 0.27 | 0.27 | 1.010 |
| $\boldsymbol{\lambda_e}$ | Truncated normal distribution with<br>• Mean=0<br>• Standard deviation=0.5<br>• Left-truncated at 0 | 0.53 | 0.6 | 1.010 |
| $\boldsymbol{\lambda_c}$ | Truncated normal distribution with<br>• Mean=0<br>• Standard deviation=0.5<br>• Left-truncated at 0 | 0.5 | 0.5 | 1.156 |
| $\boldsymbol{log_{10}(\mu)}$ | Normal distribution with<br>• Mean=0<br>• Standard deviation=0.5 | 1.50 | 1.6 | 1.018 |
| $\boldsymbol{\delta}$ | Normal distribution with<br>• Mean=1/(5×52)<br>• Standard deviation=0.5 | 1/(5.6×52) | 1/(5.6×52) | 1.029 |

| | | | | |
|---|---|---|---|---|
| | • Left-truncated at 1/(8×52)<br>• Right-truncated at 1/52 | | | |
| $\boldsymbol{\varepsilon}$ | Truncated normal distribution with<br>• Mean=1.4<br>• Standard deviation=0.5<br>• Left-truncated at 1<br>• Right-truncated at 1.65 | 1.47 | 1.47 | 1.042 |
| $\boldsymbol{S_0}$ | Truncated normal distribution with<br>• Mean=0.01×N (2,629.6)<br>• Standard deviation=0.01×N (2,629.6)<br>• Left-truncated at 0.01×N (2,629.6)<br>• Right-truncated at 0.8×N (210,370.4) | 4,285.7 | 165,000 | 1.002 |
| $\boldsymbol{I_0}$ | Truncated normal distribution with<br>• Mean=0<br>• Standard deviation=0.005×N (1,314.8)<br>• Left-truncated at 0<br>• Right-truncated at 10,000 | 75.4 | 100 | 1.008 |
| $\boldsymbol{log_{10}(B_0)}$ | Truncated normal distribution with<br>• Mean=0<br>• Standard deviation=0.5<br>• Left-truncated at 0<br>• Right-truncated at 20 | 1 | 1 | 1.000 |
| $\boldsymbol{r}$ | Truncated normal distribution with<br>• Mean=0.01<br>• Standard deviation=0.1<br>• Left-truncated at 0.0001<br>• Right-truncated at 0.3 | 0.017 | 0.017 | 1.018 |
| $\boldsymbol{log_{10}(\psi)}$ | Normal distribution with<br>• Mean=5<br>• Standard deviation=1 | -2.47 | -2.47 | 1.016 |

The priors on the parameters, the two sets of starting values for chains 1 and 2, and Rhat are presented in Table S4. Rhat was below 1.1 for every parameter except $\boldsymbol{\lambda_c}$. This likely reflects some identifiability issues because $\boldsymbol{\lambda_c}$ is used in a linear combination with other parameters to modulate the contamination of the environment ($\mu\big(1 + \lambda_c f(rain_t)\big)$). Besides, the density plots of the posterior distribution of $\alpha_2$ and ε also suggested identifiability issues, despite their respective Rhat, because ($\varepsilon - e^{\alpha_2}$) represents the baseline net bacterial decay without the influence of environmental drivers (Figure S6). Despite the limited ability of our model to identify those parameters individually, the groups of parameters they are part of ($\mu\big(1 + \lambda_c f(rain_t)\big)$, ($\varepsilon - e^{\alpha_2}$), and ($\varepsilon - \varphi_t$)) remained identifiable (see below).

The estimates of Rhat and the density plots of the posterior distributions indicated that our model reached convergence for all the other parameters (Figures S6, S7, and S8).

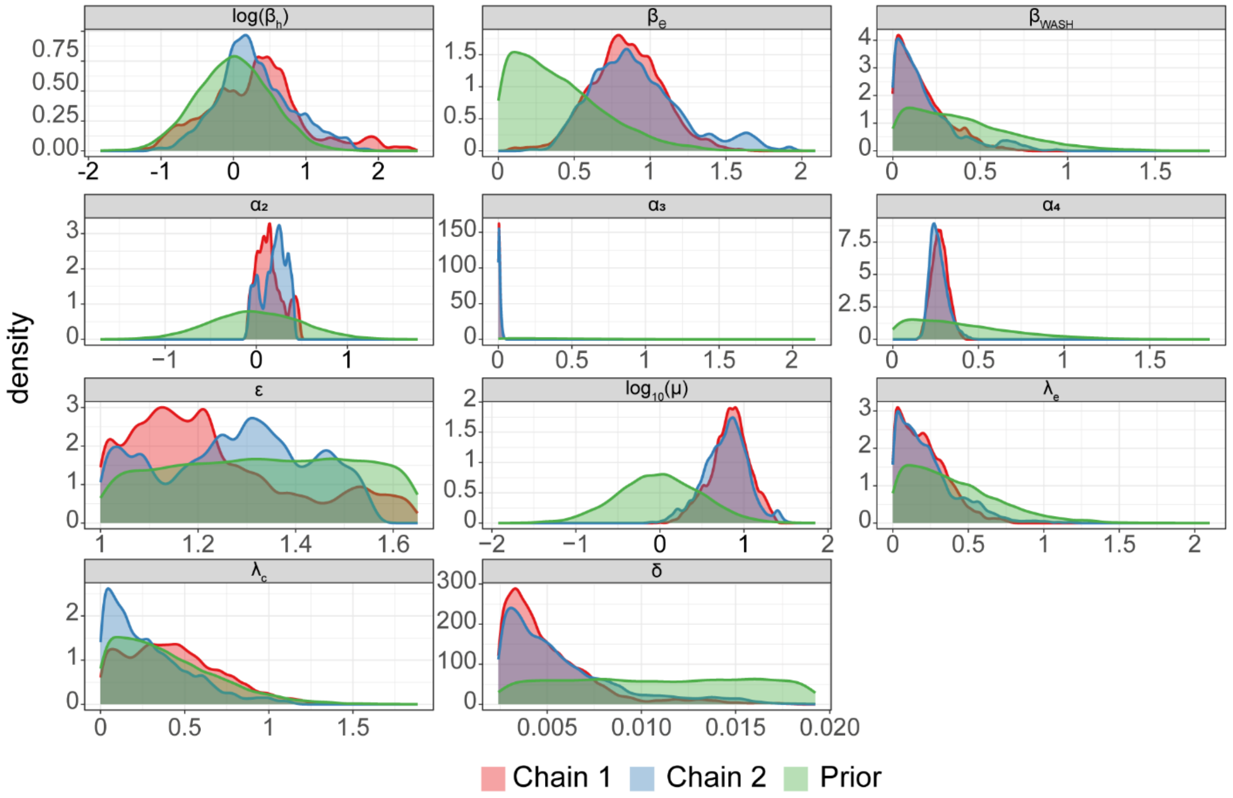


*Figure S6:* Density plots of the samples from the posterior distribution of the parameters defining cholera transmission in our model ($log\ (\beta_h)$, $\beta_e$, $\beta_{WASH}$, $\alpha_2$, $\alpha_3$, $\alpha_4$, $\mu$, $\varepsilon$, $\delta$, $\lambda_e$, $\lambda_c$) in the two chains (red for chain1 and blue for chain 2) and the corresponding prior distribution (green)

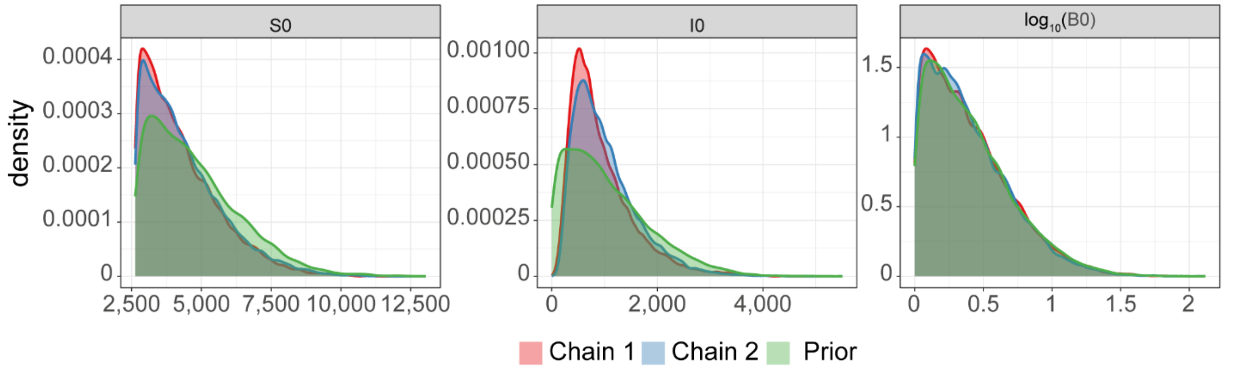


*Figure S7:* Density plots of the samples from the posterior distribution of the initial conditions (S0, I0, and B0) in the two chains (red for chain1 and blue for chain 2) and the corresponding prior distribution (green)

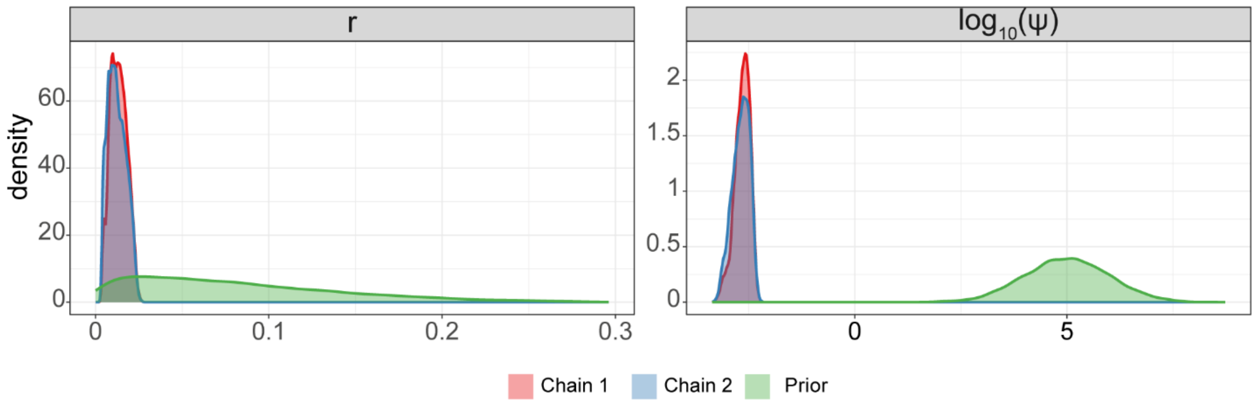


*Figure S8:* Density plots of the samples from the posterior distribution of the parameters defining the link between incident cases and reported cases (r, $\psi$) in the two chains (red for chain1 and blue for chain 2) and the corresponding prior distribution (green)

We further assessed convergence by estimating the Monte Carlo standard errors of the mean estimates of the parameters. A relative standard error (RSE) below 0.3 is commonly considered as reasonably precise, suggesting that the chain is long enough, and lack of convergence and low identifiability would tend to inflate the RSE. The RSE confirms the identifiability issue of $\boldsymbol{\alpha_2}$, with a RSE of 38.7%, but did not reflect the identifiability issue previously found with $\varepsilon$ and $\lambda_c$. All the other parameters but one had a RSE below 15%, with an RSE of 25.3% for $\log(\beta_h)$ (Figure S9).

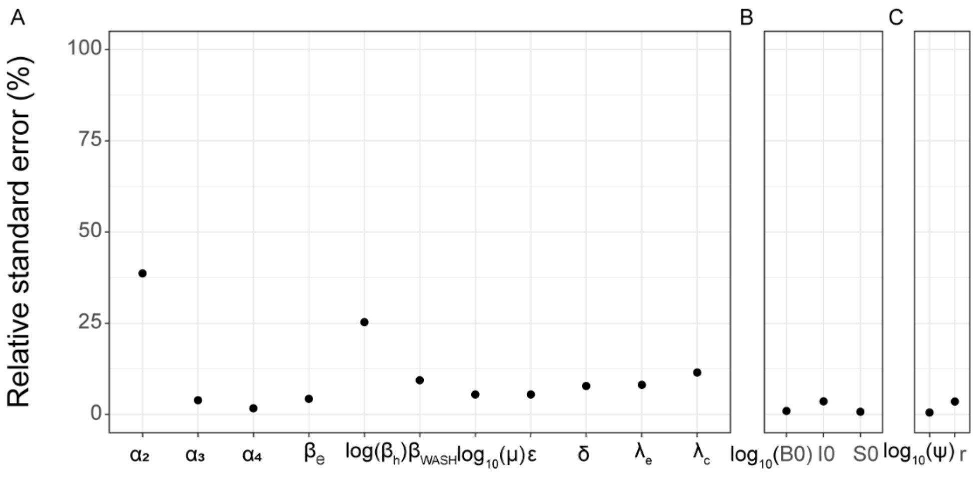


*Figure S9*: Relative standard error of the parameters defining transmission in our model (A), the initial conditions (B), and the parameters linking the incident cases to the reported cases (C)

#### Groups of parameters

The identifiability issues on $\alpha_2$, $\varepsilon$, and $\lambda_c$ still allowed us to reach convergence and identify relevant groups of parameters (Table S5 and Figure S10). This implies that the limited identifiability of $\alpha_2$, $\varepsilon$, and $\lambda_c$ did not limit our ability to estimate the impact of the intervention.

*Table S5:* Gelman and Rubin statistics (Rhat) of groups of parameters at different time points

| Group of parameters | Description | Rhat |
|---|---|---|
| $\frac{\boldsymbol{\beta_e}}{\boldsymbol{\beta_h}}$ | Ratio between the indirect and direct transmission rates | 1.000 |
| $\boldsymbol{\beta_e - \beta_{WASH}}$ | Indirect transmission rate reduced by the WASH component at the end of the study period | 1.021 |
| $(\boldsymbol{\varepsilon - e^{\alpha_2}})$ | Baseline net bacterial decay rate without the influence of environmental drivers | 1.000 |
| $\boldsymbol{\beta_e(1+\lambda_e f(rain_t))}$ for t=30 | Indirect transmission modulated by precipitation at time step 30 | 1.032 |
| $\boldsymbol{\beta_e(1+\lambda_e f(rain_t))}$ for t=60 | Indirect transmission modulated by precipitation at time step 60 | 1.038 |
| $\boldsymbol{\beta_e(1+\lambda_e f(rain_t))}$ for t=90 | Indirect transmission modulated by precipitation at time step 90 | 1.032 |
| $\boldsymbol{\mu(1+\lambda_c f(rain_t))}$ for t=30 | Environmental contamination modulated by precipitation at time step 30 | 1.024 |
| $\boldsymbol{\mu(1+\lambda_c f(rain_t))}$ for t=60 | Environmental contamination modulated by precipitation at time step 60 | 1.095 |
| $\boldsymbol{\mu(1+\lambda_c f(rain_t))}$ for t=90 | Environmental contamination modulated by precipitation at time step 90 | 1.023 |

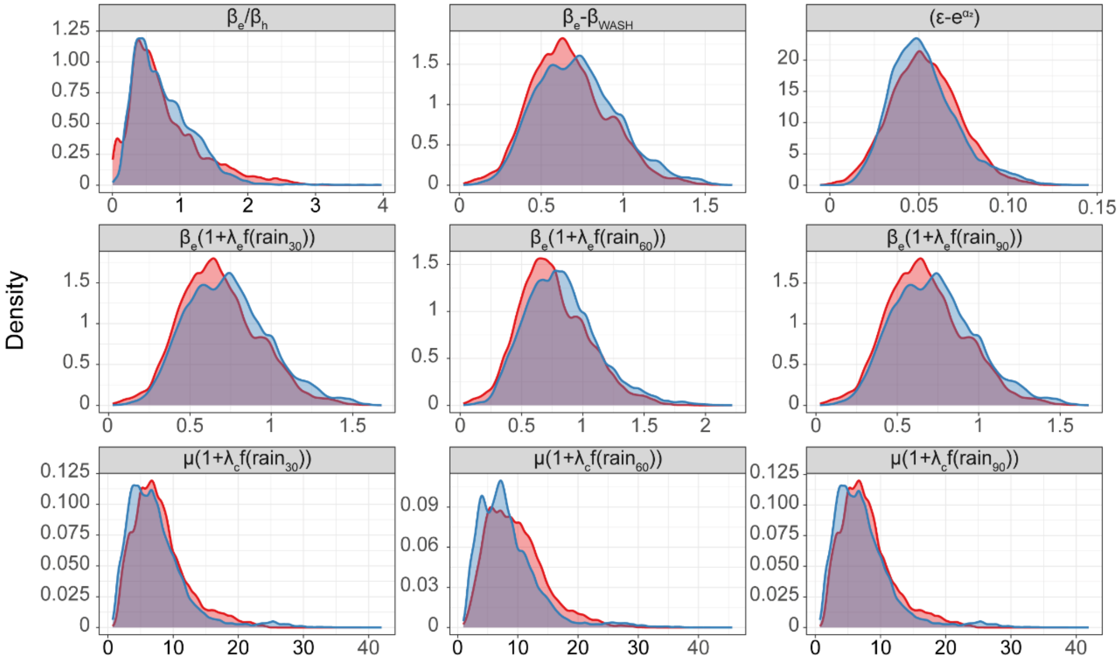


*Figure S10:* Density plots of the samples from the posterior distribution of groups of parameters influencing cholera transmission in our model in the two chains (red for chain1 and blue for chain 2)

#### Main indicators to assess the impact of the intervention

Finally, we assessed convergence of the primary indicators of interest, the ones directly used to assess the impact of the intervention: the number of infected individuals and the net bacterial growth (-net bacterial decay) at three time points, week 24 in 2014, and weeks 2 and 32 in 2015 (time steps 30, 60, and 90) (Figures S11 and S12). Those two indicators are the results of the combination of all the individual parameters. The trace plots, density plots, and Gelman and Rubin statistics all suggest that we reached convergence on those two main indicators.

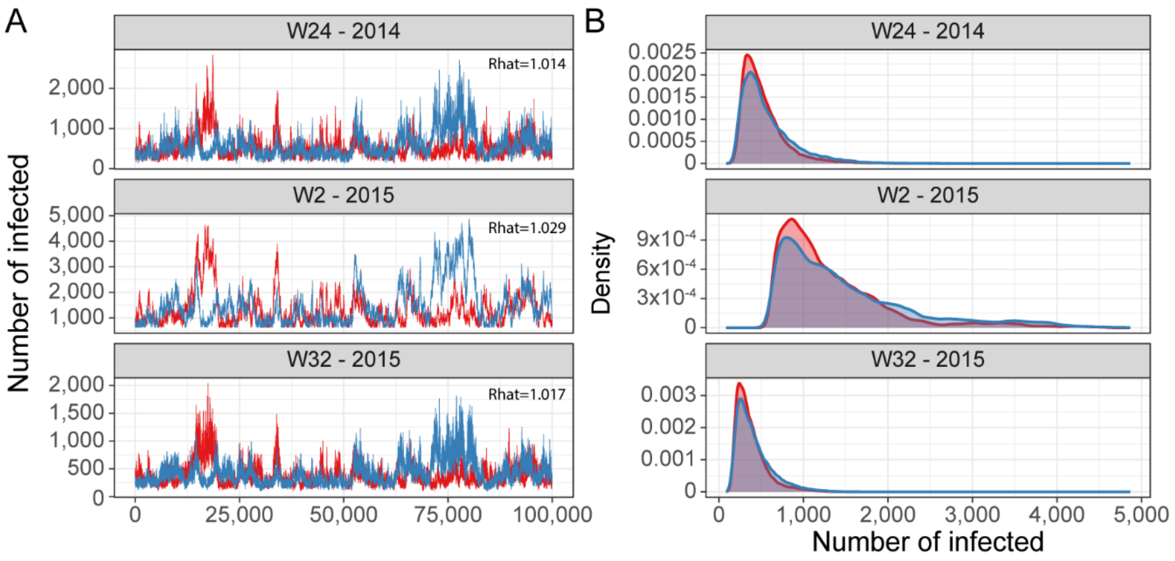


*Figure S11*: Posterior distribution of the number of infected individuals in week 24 in 2014, and weeks 2 and 32 in 2015. (A) Trace plots of the chain of the number of infected at three time points. The two chains are color coded (the first chain in red and the second in blue) and the Gelman and Rubin statistic (Rhat) is indicated. (B) Density plots of the posterior distribution of the two chains.

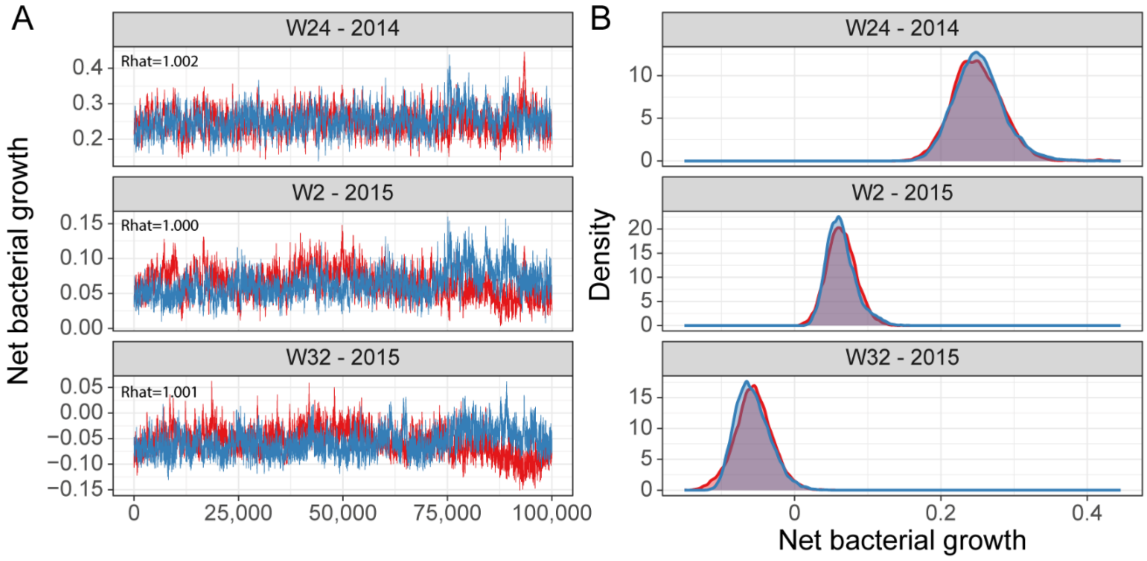


*Figure S12:* Posterior distribution of the net bacterial growth in the environmental reservoir in week 24 in 2014, and weeks 2 and 32 in 2015. (A) ) Trace plots of the chain of the net bacterial growth in the environmental reservoir at three time points.

The two chains are color coded (the first chain in red and the second in blue) and the Gelman and Rubin statistic (Rhat) is indicated. (B) Density plots of the posterior distribution of the two chains.

#### Parameter values based on the literature

The values indicated for $\varepsilon$ in Table S6 are the results of an iterative process. We used a wider range of possible values for the prior of $\varepsilon$ (1-4.2 weeks)$^{-1}$ (19,20) in preliminary fits, and then restricted its range so that we kept it as narrow as possible without clipping sampled values from its posterior distribution. The aim was to try to minimize identifiability issues with the components of $\varphi_t$. However, given the lack of identifiability after calibration, the baseline net bacterial decay rate ($\boldsymbol{\varepsilon - e^{\alpha_2}}$) should be interpreted as a whole rather than relying on estimates of its individual parameters.

*Table S6:* Parameter values based on literature

| Parameter | Description | Units | Value | References |
|---|---|---|---|---|
| $\boldsymbol{\kappa}$ | Half saturation constant | Cells/mL | $10^6$ | (19) |
| $\boldsymbol{\delta}$ | Recovery rate | Weeks$^{-1}$ | (0.71 week)$^{-1}$ | (21) |
| $\boldsymbol{\varepsilon}$ | Bacterial decay rate | Weeks$^{-1}$ | (1.05-1.65 weeks)$^{-1}$[a] | (19,20) |

a: Bacterial decay rate estimates have a wide range of values, so we clipped the parameter space by constraining its prior with some margin around values found in the literature.

#### Parameter estimates

*Table S7:* Estimates of the parameters based on the best performing model

| Parameter | Description | Units | Mean estimate (95% credible interval) |
|---|---|---|---|
| $log(\beta_h)$ | Infection rate related to interhuman transmission | Log Weeks$^{-1}$ | 0.31 (-0.99-1.83) |
| $\beta_e$ | Infection rate related to environmental exposure | Weeks$^{-1}$ | 0.84 (0.37-1.30) |
| $\beta_{WASH}$ | Reduction in infection rate related to environmental exposure to the WASH intervention | Weeks$^{-1}$ | 0.17 (0-0.46) |
| $\alpha_2$ | Baseline bacterial growth in the environment (exponential scale) | | 0.15 (-0.07-0.45) |
| $\alpha_3$ | Strength of the association between lake surface temperature variation and bacterial growth variation (exponential scale) | | 0.009 (0-0.025) |
| $\alpha_4$ | Strength of the association between chlorophyll-a variation and bacterial growth variation (exponential scale) | | 0.27 (0.18-0.36) |
| $\lambda_e$ | Strength of the amplification of the environmental exposure due to precipitation | | 0.21 (0-0.50) |
| $\lambda_c$ | Strength of the amplification of the environmental contamination due to precipitation | | 0.43 (0-0.94) |
| $\delta$ | Immunity waning rate | Weeks$^{-1}$ | 0.005 (0.002-0.011) |
| $log_{10}(\mu)$ | Contamination rate | $Log_{10}$(Cells.mL$^{-1}$.Individual$^{-1}$.Weeks$^{-1}$) | 0.83 (0.36-1.29) |
| $\varepsilon$ | Bacterial decay rate | Weeks$^{-1}$ | 1.23 (1.00-1.57) |
| $S_0$ | Susceptible individuals at time 0 | Individuals | 4,279. (2,629.7-6,968.7) |
| $I_0$ | Infectious individuals at time 0 | Individuals | 940.3 (110.6-2,165.0) |
| $log_{10}(B_0)$ | Cells at time 0 | $Log_{10}$(Cells/mL) | 0.40 (0-0.98) |
| $r$ | Reporting proportion | | 0.013 (0.004-0.022) |
| $log_{10}(\psi)$ | Overdispersion parameter | | -2.65 (-3.03—2.33) |

### Additional results

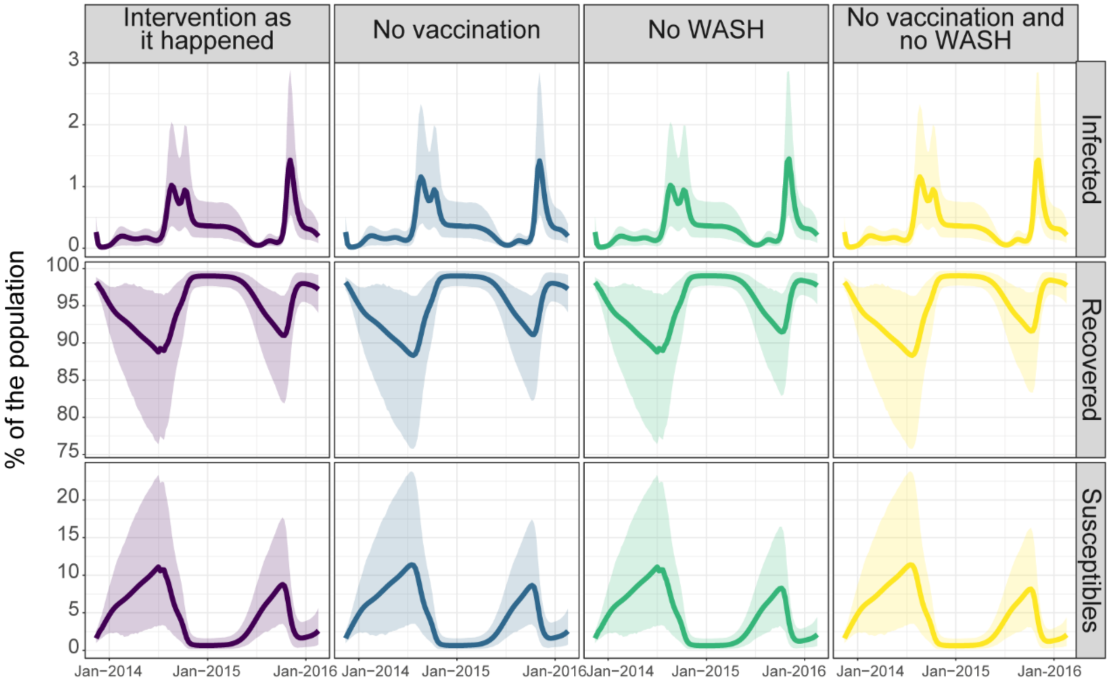


*Figure S13:* Model predictions of the mean percent of the population that was infected, recovered, and susceptible considering: the intervention as it happened of WASH and vaccination, WASH only, vaccination only, and no intervention of either vaccination or WASH from November 2013 to February 2016. The envelopes indicate the 95% credible intervals.

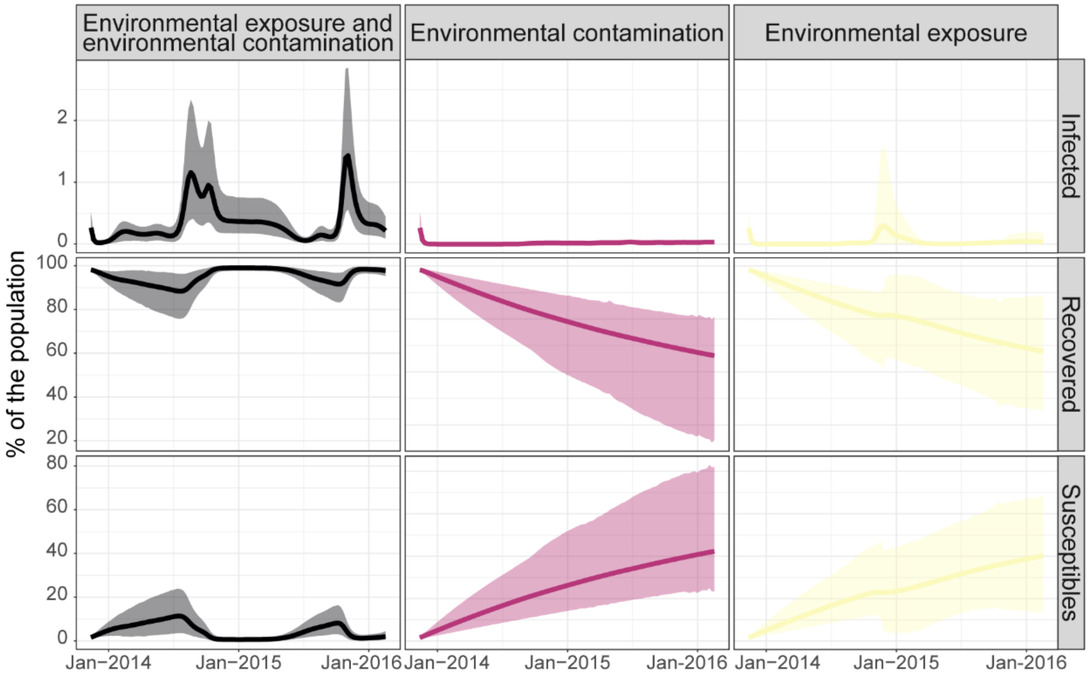


*Figure S14:* Model predictions of the mean percent of infected, recovered, and susceptible individuals in the population with environmental exposure and contamination (assuming no intervention), environmental contamination (no environmental exposure), and environmental exposure (no environmental contamination) from November 2013 to February 2016. The envelopes indicate the 95% credible intervals.

#### Sensitivity analysis: leaky vaccine

The model presented in the manuscript assume an all-or-nothing vaccine. This leads to more optimistic estimates regarding the impact of the vaccine. We then also fit an alternative of the "best performing" model with leaky vaccination keeping similar assumptions regarding immunity waning for vaccine-induced immunity to avoid adding unexplored parameters. In this alternative model we then make the simplifying assumption that immunity waning occurs at a similar rate for individuals in the recovered compartment and individuals in the two additional compartments V1 and V2. Those two compartments include individuals who respectively received one dose and 2+ doses. They can acquire the infection but transmission rates are reduced by the vaccine effectiveness of a one and two dose-regimens ($\sigma_1$ and $\sigma_2$).

The elements in blue in the differential equations below indicate the main changes compared to the "best performing" model.

$$\frac{dS}{dt} = \delta(R + V_1 + V_2) - \beta_h \frac{SI}{N} - \beta_e \frac{B}{\kappa + B}\left(1 + \lambda_e f(rain_t)\right)S - \eta_1 S - \eta_2 S$$

$$\frac{dV_1}{dt} = \eta_1 S - \left(\beta_h \frac{I}{N} + \beta_e \frac{B}{\kappa + B}\left(1 + \lambda_e f(rain_t)\right)\right)(1 - \sigma_{1_1})V_1 - \delta V_1$$

$$\frac{dV_2}{dt} = \eta_2 S - \left(\beta_h \frac{I}{N} + \beta_e \frac{B}{\kappa + B}\left(1 + \lambda_e f(rain_t)\right)\right)(1 - \sigma_2)V_2 - \delta V_2$$

$$\frac{dI}{dt} = \left(\beta_h \frac{I}{N} + \beta_e \frac{B}{\kappa + B}\left(1 + \lambda_e f(rain_t)\right)\right)(S + (1 - \sigma_1)V_1 + (1 - \sigma_2)V_2) - \gamma I$$

$$\frac{dR}{dt} = \gamma I - \delta R + \eta S$$

$$\frac{dB}{dt} = \mu\left(1 + \lambda_c f(rain_t)\right)I - B(\varepsilon - \varphi_t)$$

with

$$\varphi_t = e^{\alpha_2 + \alpha_3 sst_t + \alpha_4 chlor_t}$$

$$\eta_1 = \theta VC_{1t}$$

$$\eta_2 = \theta VC_{2t}$$

$$f(rain_t) = \frac{rain_t}{max(rain_t)}$$

The model assuming a leaky vaccine provides alternative, less optimistic, estimates of the impact of the vaccination campaign, because it allows vaccinated individuals to contribute to transmission. This likely also influences the estimates regarding the impact of the WASH component.

This model structure had a comparable model fit, with WAIC that did not significantly differ from value of the model presented in the manuscript (WAIC=792.7 with a standard error of 31.7 vs 7WAIC=87.8 with a standard error of 31.3) (Figure S15A). The model estimates of how the proportions of susceptible, infected and recovered individuals changed over time were very similar to the model assuming an all-or-nothing vaccine (Figure S15B).

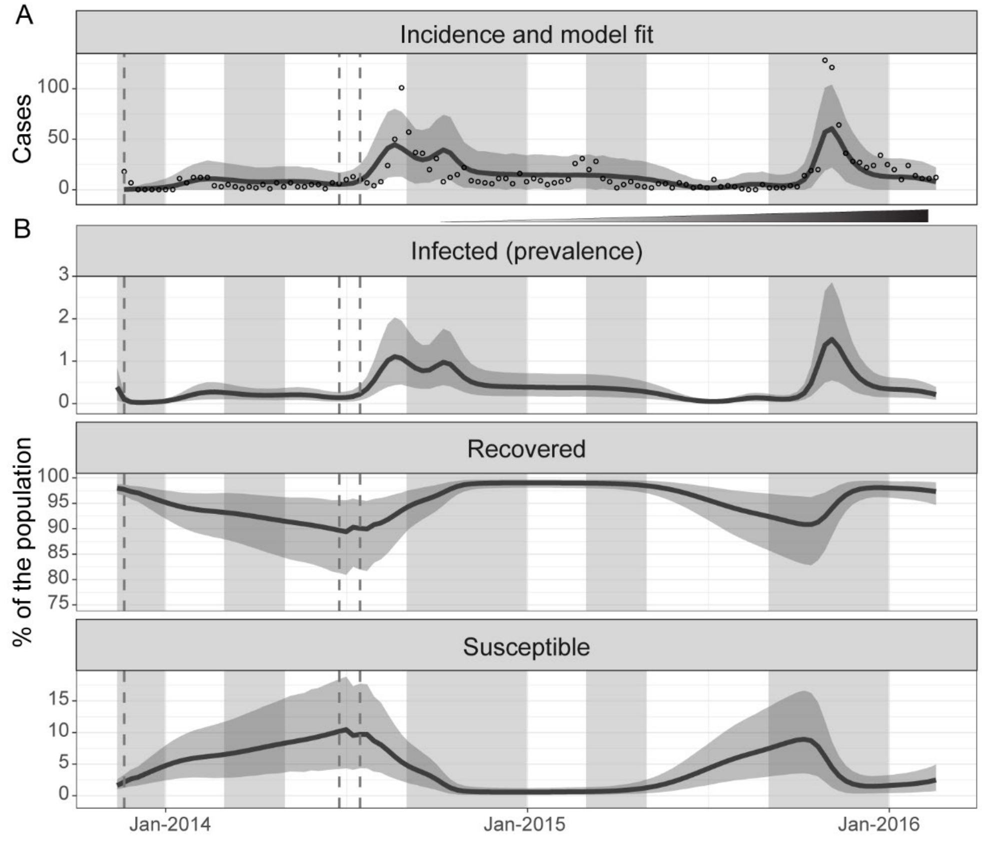


*Figure S15: Incident cases, model fit, and variation of the percentage of infected, recovered, and susceptible over time when assuming a leaky vaccine. (A) Weekly reported suspected cholera cases residing in the city of Kalemie (empty circles) from November 2013 to February 2016 and mean model prediction of the reported weekly cholera cases (dark line) and its 95% credible interval (grey envelope) (B) Mean model prediction of the percent of the population infected (prevalence), recovered, and susceptible (dark lines) and their 95% credible interval (grey envelopes) from November 2013 to February 2016. Typical rainy seasons are shaded in grey, the timing of the distribution of vaccine doses in vertical dashed grey lines, and the*

*incremental implementation of the improvements in water and sanitation is indicated by the thickening and darkening horizontal line.*

Assuming a leaky vaccine led to estimates of a smaller impact of the vaccination. The scenario without vaccination (WASH only), without WASH (vaccination only), and without vaccination and WASH led to an average increase of 293 (mean: 293.2, 95%CrI: 60.9-735.6), 1,677 (mean: 1,677.4, 95%CrI: 0.3-4,864.1), and 1,952 cases (mean: 1,951.9, 95rI: 83.3-5,277.9) respectively (Figure S16).

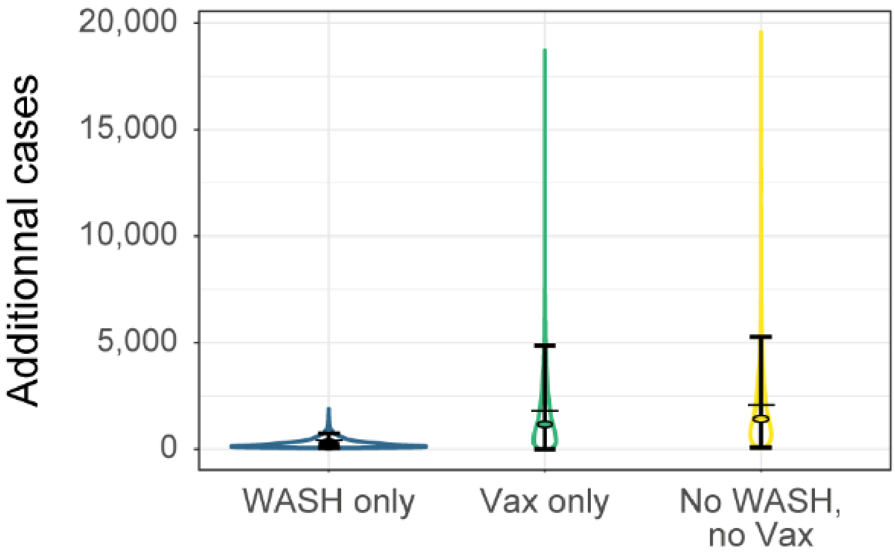


*Figure S16: : Estimated impact of the components of the intervention when assuming a leaky vaccine. Violin plots of numbers of additional cholera cases at the end of the study period with WASH only (blue), vaccination only (green), or no intervention (no WASH and no vaccination) (yellow) compared to the intervention as it happened of WASH and vaccination. The error bars, the filled circles, and the horizontal bars indicate the 95% credible interval, the medians, and the means respectively.*

#### Parameter estimates when assuming a leaky vaccine

*Table S8: Estimates of the parameters based on the modification of the best performing model assuming a leaky vaccine*

| Parameter | Description | Units | Mean estimate (95% credible interval) |
|---|---|---|---|
| $log(\beta_h)$ | Infection rate related to interhuman transmission | Log Weeks$^{-1}$ | 0.08 (-0.83-1.18) |
| $\beta_e$ | Infection rate related to environmental exposure | Weeks$^{-1}$ | 0.96 (0.46-1.57) |
| $\beta_{WASH}$ | Reduction in infection rate related to environmental exposure to the WASH intervention | Weeks$^{-1}$ | 0.22 (0-0.56) |
| $\alpha_2$ | Baseline bacterial growth in the environment (exponential scale) | | 0.34 (-0.01-0.49) |
| $\alpha_3$ | Strength of the association between lake surface temperature variation and bacterial growth variation (exponential scale) | | 0.009 (0-0.024) |
| $\alpha_4$ | Strength of the association between chlorophyll-a variation and bacterial growth variation (exponential scale) | | 0.23 (0.16-0.32) |
| $\lambda_e$ | Strength of the amplification of the environmental exposure due to precipitation | | 0.23 (0-0.60) |
| $\lambda_c$ | Strength of the amplification of the environmental contamination due to precipitation | | 0.30 (0-0.75) |
| $\delta$ | Immunity waning rate | Weeks$^{-1}$ | 0.005 (0.002-0.010) |
| $log_{10}(\mu)$ | Contamination rate | $Log_{10}$(Cells.mL$^{-1}$.Individual$^{-1}$.Weeks$^{-1}$) | 0.76 (0.37-1.19) |
| $\varepsilon$ | Bacterial decay rate | Weeks$^{-1}$ | 1.45 (1.05-1.65) |
| $S_0$ | Susceptible individuals at time 0 | Individuals | 4,275.6 (2,629.7-9,898.5) |
| $I_0$ | Infectious individuals at time 0 | Individuals | 1,020.2 (174.4-2,189.9) |
| $log_{10}(B_0)$ | Cells at time 0 | $Log_{10}$(Cells/mL) | 0.40 (0-0.98) |
| $r$ | Reporting proportion | | 0.122 (0.005-0.02) |
| $log_{10}(\psi)$ | Overdispersion parameter | | -2.70 (-3.03—2.39) |

### References


1. Page AL, Alberti KP, Mondonge V, Rauzier J, Quilici ML, Guerin PJ. Evaluation of a rapid test for the diagnosis of cholera in the absence of a gold standard. PLoS ONE. 2012;7(5):e37360.

2. Nadri J, Sauvageot D, Njanpop-Lafourcade BM, Baltazar CS, Banla Kere A, Bwire G, et al. Sensitivity, Specificity, and Public-Health Utility of Clinical Case Definitions Based on the Signs and Symptoms of Cholera in Africa. Am J Trop Med Hyg. 2018 Apr;98(4):1021–30.

3. Harris JB, LaRocque RC, Qadri F, Ryan ET, Calderwood SB. Cholera. The Lancet. 2012 Jun 30;379(9835):2466–76.

4. Massing LA, Aboubakar S, Blake A, Page AL, Cohuet S, Ngandwe A, et al. Highly targeted cholera vaccination campaigns in urban setting are feasible: The experience in Kalemie, Democratic Republic of Congo. PLOS Neglected Tropical Diseases. 2018 May 7;12(5):e0006369.

5. Luquero FJ, Grout L, Ciglenecki I, Sakoba K, Traore B, Heile M, et al. Use of Vibrio cholerae vaccine in an outbreak in Guinea. N Engl J Med. 2014 May 29;370(22):2111–20.

6. Ferreras E, Chizema-Kawesha E, Blake A, Chewe O, Mwaba J, Zulu G, et al. Single-Dose Cholera Vaccine in Response to an Outbreak in Zambia. N Engl J Med. 2018 08;378(6):577–9.

7. Sutton P, Roberts D, Elvidge C, Baugh K. Census from Heaven: An estimate of the global human population using night-time satellite imagery. International Journal of Remote Sensing. 2001 Jan 1;22(16):3061–76.

8. Bharti N, Djibo A, Tatem AJ, Grenfell BT, Ferrari MJ. Measuring populations to improve vaccination coverage. Sci Rep [Internet]. 2016 Oct 5 [cited 2019 Aug 5];5. Available from: https://www.ncbi.nlm.nih.gov/pmc/articles/PMC5050518/

9. Bharti N, Tatem AJ. Fluctuations in anthropogenic nighttime lights from satellite imagery for five cities in Niger and Nigeria. Sci Data [Internet]. 2018 Nov 13 [cited 2019 Jul 30];5. Available from: https://www.ncbi.nlm.nih.gov/pmc/articles/PMC6233255/

10. Stathakis D, Baltas P. Seasonal population estimates based on night-time lights. Computers, Environment and Urban Systems. 2018 Mar 1;68:133–41.

11. Bharti N, Lu X, Bengtsson L, Wetter E, Tatem AJ. Remotely measuring populations during a crisis by overlaying two data sources. Int Health. 2015 Mar;7(2):90–8.

12. Bharti N, Tatem AJ, Ferrari MJ, Grais RF, Djibo A, Grenfell BT. Explaining Seasonal Fluctuations of Measles in Niger Using Nighttime Lights Imagery. Science. 2011 Dec 9;334(6061):1424–7.

13. Elvidge CD, Baugh K, Zhizhin M, Hsu FC, Ghosh T. VIIRS night-time lights. International Journal of Remote Sensing. 2017 Nov 2;38(21):5860–79.

14. NASA O. Moderate-resolution Imaging Spectroradiometer (MODIS) Aqua Ocean Color Data; 2014 Reprocessing. NASA OB. DAAC. Greenbelt, MD, USA: NASA Goddard Space Flight Center, Ocean Ecology Laboratory, Ocean Biology Processing Group. 2014.

15. Bergamino N, Horion S, Stenuite S, Cornet Y, Loiselle S, Plisnier PD, et al. Spatio-temporal dynamics of phytoplankton and primary production in Lake Tanganyika using a MODIS based bio-optical time series. Remote Sensing of Environment. 2010 Apr 15;114(4):772–80.

16. Sheffield J, Goteti G, Wood EF. Development of a 50-Year High-Resolution Global Dataset of Meteorological Forcings for Land Surface Modeling. J Climate. 2006 Jul 1;19(13):3088–111.

17. Lemaitre J, Pasetto D, Perez-Saez J, Sciarra C, Wamala JF, Rinaldo A. Rainfall as a driver of epidemic cholera: Comparative model assessments of the effect of intra-seasonal precipitation events. Acta Tropica. 2019 Feb 1;190:235–43.

18. Watanabe S, Opper M. Asymptotic equivalence of Bayes cross validation and widely applicable information criterion in singular learning theory. Journal of machine learning research. 2010;11(12).

19. Codeço CT. Endemic and epidemic dynamics of cholera: the role of the aquatic reservoir. BMC Infectious Diseases. 2001 Feb 2;1(1):1.

20. Dimitrov DT, Troeger C, Halloran ME, Longini IM, Chao DL. Comparative Effectiveness of Different Strategies of Oral Cholera Vaccination in Bangladesh: A Modeling Study. PLOS Neglected Tropical Diseases. 2014 Dec 4;8(12):e3343.

21. Sack DA, Sack RB, Nair GB, Siddique AK. Cholera. Lancet. 2004 Jan 17;363(9404):223–33.
